# A national atlas of county-level decarceration mechanisms and their association with mortality across the United States

**DOI:** 10.64898/2026.04.02.26349309

**Authors:** Yiran E. Liu, Beier Li, Joshua L. Warren, Gregg S. Gonsalves, Emily A. Wang

**Author notes:** Corresponding author: Yiran E. Liu, PhD, MS, 60 Temple Street, Suite 5C, New Haven, CT 06511.

## Abstract

Decarceration, the process of reducing incarceration rates, is increasingly proposed as a strategy to improve population health. Yet, its health effects remain understudied and may depend on how and where decarceration occurs. We developed a national decarceration “atlas” to systematically characterize local mechanisms of decarceration and examine their associations with population-level mortality across 2,870 counties in the United States between 1999-2019. Using longitudinal county-level data, we identified four operational types: reduced pretrial detention, reduced jail time, reduced prison admissions, and reduced prison time. We validated this classification against documented case studies of local and state decarceration. While most counties experienced at least one decarceration type during the period, declines were typically isolated to one part of one system and modest in magnitude, with median reductions of 19% to 38% ten years post-onset. Of the four types, reduced pretrial detention and reduced prison admissions – the two front-end strategies – were associated with 3.4% (95% CI, 0.7-6.1) and 1.9% (95% CI, 0.2-3.5) lower all-cause mortality, respectively, in large central metropolitan counties. By contrast, associations were consistent with the null for smaller urban and rural counties, and for mechanisms reducing the length of jail or prison incarceration. These findings indicate that decarceration is a heterogeneous phenomenon that is not universally health-promoting. Strategies to prevent incarceration may be more impactful for population health than those that shorten time served, but this pattern likely depends on local context. Further work is needed to understand the conditions under which decarceration advances public health.

## INTRODUCTION

The United States (U.S.) has one of the highest incarceration rates in the world, with documented adverse, unequal effects on individual, family, and community health^1^. Decarceration, the process of reducing incarceration rates, is increasingly viewed as an opportunity to improve population health and health equity^2^. Since 2007, incarceration rates have declined modestly nationwide, driven by considerable reductions in select states and jurisdictions^3^. Yet, evidence on the community health effects of these declines remains limited.

Decarceration may not simply reverse the health harms of mass incarceration. Decades of concentrated incarceration have reshaped the social, economic, and political conditions of communities in ways that may not be quickly or easily undone^4^. Moreover, unmet health needs among people with criminal legal involvement can make incarceration a point of contact for health services, even amid concerns about accessibility and quality of care in custody^5–9^. Whether decarceration promotes population health is likely context-dependent, with myriad drivers of heterogeneity.

Prior literature supports this complexity. A handful of studies report associations between county-level incarceration rates and mortality^10–12^. However, these estimates come from periods of predominantly rising incarceration and assume, rather than test, that increases and decreases have equal and opposite effects. Direct study of decarceration has been limited. Single-state policy evaluations have found null effects of bail reform and drug decriminalization on violence^13,14^ and fatal overdose^15,16^, respectively. Meanwhile, COVID-19-era decarceration has been linked to reduced infection rates^17,18^ alongside health and social service disruptions for people with criminal-legal involvement^8,19^. Together, this work indicates that decarceration may have mixed and possible adverse health effects in some settings, highlighting the need to better understand what drives this heterogeneity and identify strategies for health-promoting decarceration.

A major impediment to this line of inquiry is the lack of standardized measures for how decarceration has occurred across jurisdictions. There is no comprehensive database of decarceral reforms. Even if it were possible to systematically document all criminal legal reforms across geographic levels (i.e., federal, state, local) over time, such a database would have substantial limitations. Reforms exhibit considerable variation across jurisdictions^20,21^; legislation does not always translate into reduced incarceration^22–24^; and implementation of laws varies across settings^25,26^. Moreover, decarceration can occur through local, *de facto* processes not captured in legislation, such as budget cuts, investments in diversion programs, or shifts in judicial, prosecutorial, or policing practices^27–29^.

A complementary approach to policy surveillance that overcomes these challenges involves characterizing decarceration based on observed reductions, with decomposition of different operational mechanisms across jail and prison systems. Publicly available longitudinal administrative data include multiple measures aggregated to the county level, enabling distinction of “front-end” reductions that divert people away from jail or prison and “back-end” reductions in time served for people already incarcerated. While each of these processes can arise through diverse *de jure* and *de facto* pathways, this approach provides a scalable framework to examine how the operational mechanism of decarceration may differentially affect health, and how effects may vary across settings.

In this study, we introduce a typology of decarceration mechanisms spanning jail and prison systems and provide the first nationwide, county-level investigation of their distribution and health effects in more than 2,800 U.S. counties between 1999 and 2019. We validate classifications against well-documented instances of real-world decarceration and characterize the frequency, timing, magnitude, and dynamics of each decarceration type. We then estimate county-level associations between each decarceration type and all-cause mortality, assessing effect modification by county urbanicity. We hypothesized that front-end reductions, reflecting upstream prevention and earlier “off ramps”, would be more strongly associated with reduced mortality than back-end reductions. We additionally hypothesized that associations would be more pronounced in urban counties, where there is greater health and social service infrastructure than in rural areas to support the needs of people remaining in or returning to the community^30,31^.

## METHODS

### Data sources and pre-processing

We use county-level incarceration data from the Vera Institute of Justice from 1999 through 2019, the only period for which all of the following measures were available yearly at the county level: pretrial jail population, total jail population, prison admissions, and prison population^32^. County resident population aged 15-64 was obtained from the U.S. Census Bureau to approximate the population at greatest risk of incarceration. County-level prison measures were based not on the county of imprisonment (i.e., where a prison is located), but on the county of court commitment (i.e., where the legal process takes place). This is often, but not always, a person’s county of residence pre-incarceration^33^. Prison incarceration measures should therefore be interpreted as indicators of local criminal-legal practices (i.e., court and prosecutorial decision-making). We included counties with jail and/or prison data for at least 15 timepoints, with gaps of no more than three consecutive years. We excluded states with unified prison and jail systems, for which county-level measures were unavailable or incomplete in the Vera data^34^. Urbanicity was classified using collapsed categories from the National Center for Health Statistics (NCHS): 1) large central metro, 2) large fringe metro, 3) small-medium metro, and 4) rural. Further details are provided in the **Appendix**.

We obtained restricted-use county-level mortality data from NCHS. We additionally acquired county-level covariate data on demographics, poverty, median household income, education, unemployment, and crime from the U.S. Census Bureau, the Bureau of Labor Statistics, and the Federal Bureau of Investigation’s Uniform Crime Reporting Program. For covariates not measured annually or not available over the full study period, we used linear interpolation, last observation carried forward, or next observation carried backward (**Appendix**).

### Classification of decarceration types

We identified four types of decarceration distinguishable using the four incarceration measures: 1) reduced pretrial detention, 2) reduced jail time, 3) reduced prison admissions, and 4) reduced prison time. **Table 1** summarizes the criteria and real-world examples for each type. Each type was defined by a decreasing trend in a target incarceration measure; some types were additionally conditional on trends in another measure. For example, reduced jail time was defined by the total jail population decreasing more steeply than the pretrial jail population.

**Table 1.** Defining criteria and real-world examples of decarceration types.

| Type | Real-world examples | Pretrial jail population rate | Total jail population rate | Prison admissions rate | Prison population rate |
| --- | --- | --- | --- | --- | --- |
| **Reduced pretrial detention** | Bail reductions, cite-and-release practices, pre-trial diversion, alternative crisis response models | Decreasing | Any | Any | Any |
| **Reduced jail time** | Diversion, de-prosecution or reduced sentences for minor offenses, capacity-based releases | Non-decreasing, or decreasing less steeply* | Decreasing | Any | Any |
| **Reduced prison admissions** | Felony-to-misdemeanor reclassifications, diversion, reduced incarceration for supervision violations | Any | Any | Decreasing | Any |
| **Reduced prison time** | Earned time/good time credits, second-look resentencing, expanded parole eligibility, elimination of mandatory minimums | Any | Any | Non-decreasing | Decreasing |

\*Decreasing less steeply than the total jail population rate.

### Incarceration trend ascertainment

To identify decreasing trends, we applied piecewise negative binomial regression^35^ separately for each county and incarceration measure. The incarceration measure was the response variable, year the explanatory variable, with the log resident population aged 15-64 as an offset. We fit negative binomial regression models with zero, one, or two breakpoints (continuous between segments)^36^, estimated algorithmically^35^, and selected the model with the lowest Akaike information criterion (AIC). We extracted segment slopes and p-values to characterize trend direction and uncertainty. P-values were adjusted for multiple comparisons using the Benjamini-Hochberg method. We classified segments as decreasing if they had a negative slope and an adjusted p-value ≤ 0.05. To reduce edge effects from truncated observation windows, we did not classify segments as decreasing if they started at the first timepoint and lasted ≤4 years, or if they started within four years of the final timepoint. We excluded segments with cumulative declines of fewer than 10 individuals, to avoid labeling negligible fluctuations as decarceration. All remaining segments were classified as non-decreasing.

Segment trends were then mapped to calendar years. Using criteria in **Table 1**, we assigned a binary, time-varying indicator for each decarceration type in each county-year. **Figure S1** and **Table S1** illustrate this process for an example county.

### Case study validation

We assessed construct validity by comparing results against a set of well-documented decarceration case studies. Because no gold-standard dataset of county-level decarceration exists, our objective was not to assess sensitivity or specificity, but to evaluate whether the method identifies expected decarceration types in settings where the timing and underlying mechanisms of change are well established. Case studies were considered supportive of construct validity if the dominant decarceration type(s) identified by our approach aligned with documented mechanisms and occurred in the expected period.

We selected the following eight case studies spanning: all four decarceration types; diverse real-world mechanisms (e.g. statutory reforms, court orders, fiscal constraints, diversion programs, de facto shifts in practice); and heterogeneous geography, urbanicity and political context. Details are in the **Appendix**.

1. Public Safety Realignment in California, 2011: responsibility for lower-level offenses shifted from state prisons to counties, leading to sharp reduction in prison admissions
2. Justice Reinvestment Act in North Carolina, 2011: reduced prison time for supervision violations (shorter-term imprisonment instead of full revocation)
3. Voluntary (2006) and presumptive (2013) sentencing guidelines in Alabama: standardized prison sentences and increased use of non-prison alternatives (reduced prison time and reduced prison admissions)
4. Prisoner Reentry Initiative in Michigan, 2003: expedited prison releases (reduced prison time) through increased parole capacity and approval rates
5. Recession- and tax reform-induced shifts in policing practice and pretrial reform in Broward County, Florida (large fringe metro), 2007: closure of jail wings, decreased jail bookings, expanded pretrial release program
6. Budget shortages and reduced jail capacity in Lane County (small-medium metro), Oregon, 2002-2013: early jail releases and *de facto* de-prosecution of lower-level offenses (reduced jail time) until voters passed levy to fund more jail beds
7. Drug court and response to methamphetamine epidemic in Coles County (rural), Illinois, 2003-2005: treatment-based diversion and precursor control (reduced pretrial detention)
8. Hurricane Katrina and a cascade of jail reforms in Orleans Parish (large central metro), Louisiana, 2005-2018: reduced jail capacity, federal monitoring, and pretrial reform (reduced jail time and reduced pretrial detention)

### Magnitude and archetypal trajectories of decarceration

We identified counties experiencing each decarceration type at least once during the study period and aligned counties by the first year in which the decarceration type was classified (year 0). For counties exhibiting the same type in multiple non-contiguous intervals, the first episode was used to determine the onset year. For magnitude, we calculated the percent change in the target incarceration measure each year post-decarceration onset, relative to its level at onset. Archetypal trajectories were constructed by calculating the median rate of each incarceration measure across counties, standardized by the resident population aged 15-64, in years before and after onset.

### Analysis of decarceration and mortality

We used multivariable Poisson regression to model the association between each decarceration type and all-cause mortality. The exposure was the binary presence or absence of a decarceration type in a given county-year. The outcome was the number of deaths from any cause, with a log population offset and a one-year lead to ensure correct temporal order (i.e. exposure and covariates precede the outcome). Models included county fixed effects to account for time-invariant county-level characteristics (e.g., urbanicity, baseline incarceration rate) and year fixed effects to account for temporal shocks common to all counties (e.g., nationwide recessions, opioid epidemic waves). Estimates therefore reflect within-county associations between changes in decarceration status and subsequent mortality. We chose to use fixed rather than random effects for counties to avoid potential bias from non-independence of county random effects (i.e., counties that decarcerate may systematically differ in baseline mortality). While recent methodological work has identified potential biases of the two-way fixed effects approach in some contexts^37,38^, a diagnostic suggested minimal influence of this bias in the present analysis (**Appendix**). Models additionally adjusted for time-varying county-level covariates: median age, percent Black residents, median household income, poverty, educational attainment, unemployment, and violent crime. Continuous covariates were scaled to facilitate interpretation. Standard errors were clustered at the county level to account for serial correlation within counties over time. Each decarceration type was modeled separately, with a sensitivity analysis including all four types simultaneously. We additionally assessed effect modification by county urbanicity, reporting stratum-specific estimates and ratios of rate ratios as measures of effect modification on the multiplicative scale.

## RESULTS

Analyses included a total of 2,870 counties heterogeneous in geography, urbanicity, and sociodemographic characteristics: 2,663 with jail data, 2,184 with prison data, and 1,977 with both jail and prison data (**Figure S2, Table S2**).

### Validation against documented real-world decarceration

We validated the typology against eight well-documented instances of decarceration, detailed in the **Appendix**. Across case studies, the classified decarceration types aligned with documented mechanisms and timing. For example, consistent with California’s Public Safety Realignment, which sharply reduced statewide prison admissions in 2011, we identified a sudden surge in the number of California counties classified as experiencing reduced prison admissions in 2010-2011 (**Figure S3A**). In North Carolina, implementation of the Justice Reinvestment Act reduced the prison population while expanding community supervision and increasing the use of short-term incarceration sanctions for supervision violations. Correspondingly, our approach shows an increase in the prevalence of reduced prison time and a decrease in the prevalence of reduced prison admissions in North Carolina over the corresponding timeframe (**Figure S3A**). Following sentencing guidelines in Alabama and sentencing, parole, and reentry reforms in Michigan, we observed increases in the number of counties classified as experiencing reduced prison admissions and reduced prison time (**Figure S3A**). In all cases except California, we identified substantial numbers of counties that did not experience prison decarceration following state-level reforms. Indeed, these counties continued to experience rising prison admissions and/or prison population rates, demonstrating the method’s ability to detect intra-state heterogeneity in uptake of state-level reforms (**Figure S3B**).

Our approach also classified jail decarceration arising from varied local drivers, including municipal ordinances, court-ordered reform, and fiscal constraints (**Figure S3C**). In Broward County (large fringe metropolitan, Florida), Lane County (small-medium metropolitan, Oregon), Coles County (rural, Illinois), and Orleans and St. Bernard Parishes (large central and fringe metropolitan, respectively, Louisiana), the types of jail decarceration identified by our approach were consistent with documented local changes (**Appendix**). In some cases, estimated start years preceded documented changes by 1-2 years, reflecting limitations in capturing the exact timing of abrupt changes.

### Frequency and timing of decarceration types

Among counties with both jail and prison data, 65% were classified as experiencing at least one decarceration type during the study period. Reduced prison admissions was the most common type, occurring in about 33% of counties, followed by reduced prison time (29%), reduced jail time (25%), and reduced pretrial detention (25%). Frequency of each decarceration type varied markedly by urbanicity. For instance, reduced prison admissions occurred in 90% of large central metropolitan counties but only 23% of rural counties (**Figure 1**). Frequency also varied considerably by state: for example, between 5% to 88% of counties within a state experienced reduced prison admissions during the period (**Figure 1, Figure S4**). Within states, counties exhibited substantial heterogeneity in whether and how jail and prison decarceration occurred (**Figures S5-S6**).

**Figure 1.**
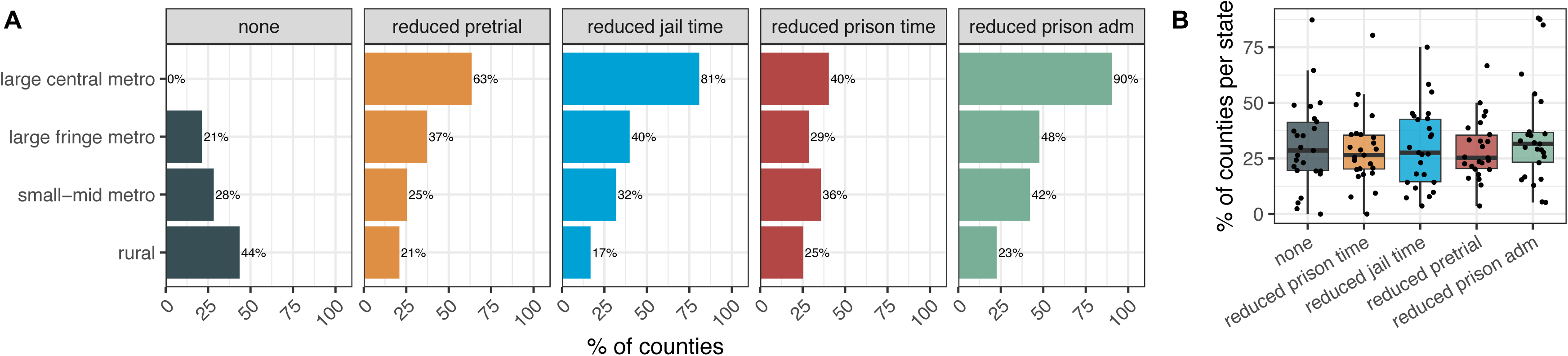
Frequency of each decarceration type by urbanicity and state. **A)** Bars show the percent of counties in each urbanicity category that exhibited each decarceration type at least once during the study period. **B)** Boxplots show the distribution, across states, of the percent of counties per state that exhibited each decarceration type at least once during the study period. Underlying points (states) are jittered horizontally. In both A and B, only counties with both jail and prison incarceration data are included to avoid misclassifying lack of data as lack of decarceration. In B, only states with data for at least 80% of counties are shown.

Regarding timing, decarceration expanded substantially during the 2000s, with the proportion of counties experiencing any decarceration increasing from 21% in 1999 to 48% by 2008 and leveling off thereafter (**Figure 2**). The number of counties experiencing multiple types concurrently also peaked near the middle of the period, reaching more than 300 counties by 2009 (**Figure S7**). The most frequent combinations were reduced jail time and reduced prison admissions, followed by reduced pretrial detention and reduced prison admissions (**Figure S7**).

**Figure 2.**
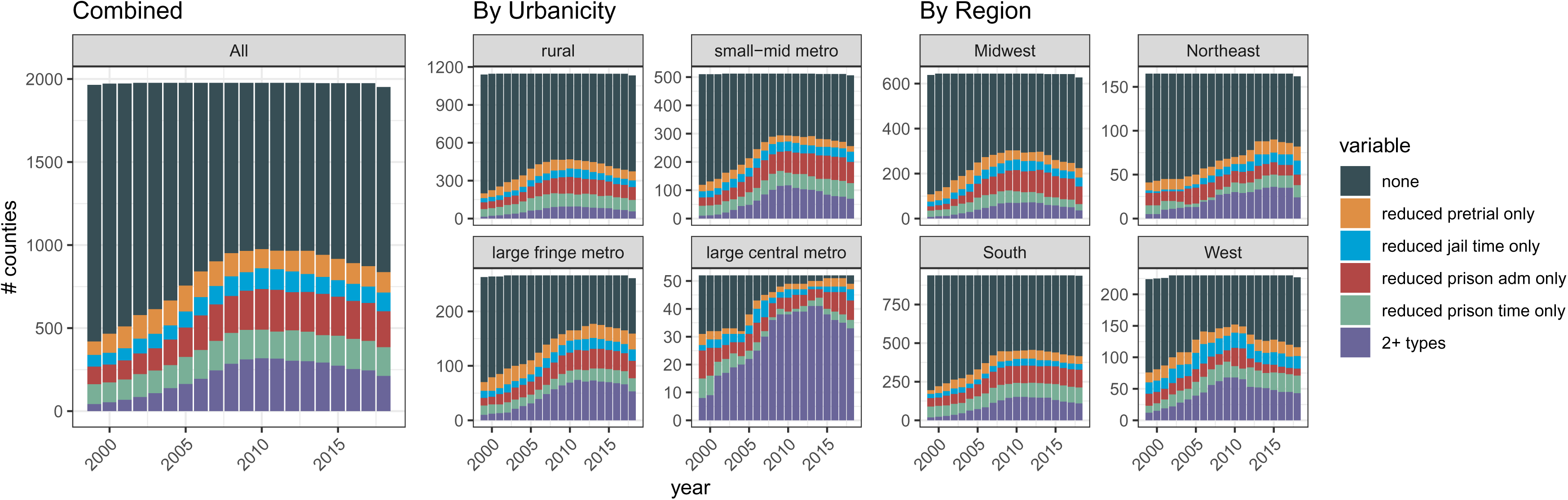
Frequency of each decarceration type over time, by county urbanicity and region. Only counties with both jail and prison incarceration data are shown.

Timing also varied by urbanicity and region (**Figure 2**). For rural and small-midsize metropolitan counties, the frequency of decarceration peaked around the middle of the period, while in large central and fringe metropolitan counties, decarceration continued to expand into the 2010s. Most large central metropolitan counties were experiencing multiple decarceration types by 2007, and nearly all maintained at least one type through the end of the period. By region, the frequency of decarceration grew across all regions in the 2000s, then leveled off or declined in the Midwest, South, and West, while continuing to grow in the Northeast through the end of the period.

### Magnitude of decarceration

We next examined the magnitude of decline for each decarceration type, relative to levels at decarceration onset **(Figure S8)**. At ten years post-onset, median declines were 37% (IQR: 22-54) for reduced pretrial detention, 24% (IQR 13-36) for reduced jail time, 38% (IQR 23-51) for reduced prison admissions, and 19% (IQR 9-31) for reduced prison time. Across types, declines were steepest in the first five to six years and leveled off thereafter. Declines were rarely reversed, with most counties maintaining lower levels ten years after onset.

### Dynamics of decarceration types

We constructed “archetypal” trajectories of all four incarceration measures in years before and after onset of each decarceration type (**Figure 3**). The number of counties contributing data for each year pre- and post-onset is shown in **Figure S9**. Trajectories post-onset were consistent with classification criteria (e.g., reduced jail time involves a decreasing total jail population rate amid a relatively stable pretrial population rate). However, pre-onset trajectories reveal that decreases generally followed increases in the same measure rather than starting from a steady state. For example, under reduced pretrial detention, pretrial rates rose before onset and declined after (**Figure 3**). As such, declines were modest compared to levels five years pre-onset.

**Figure 3.**
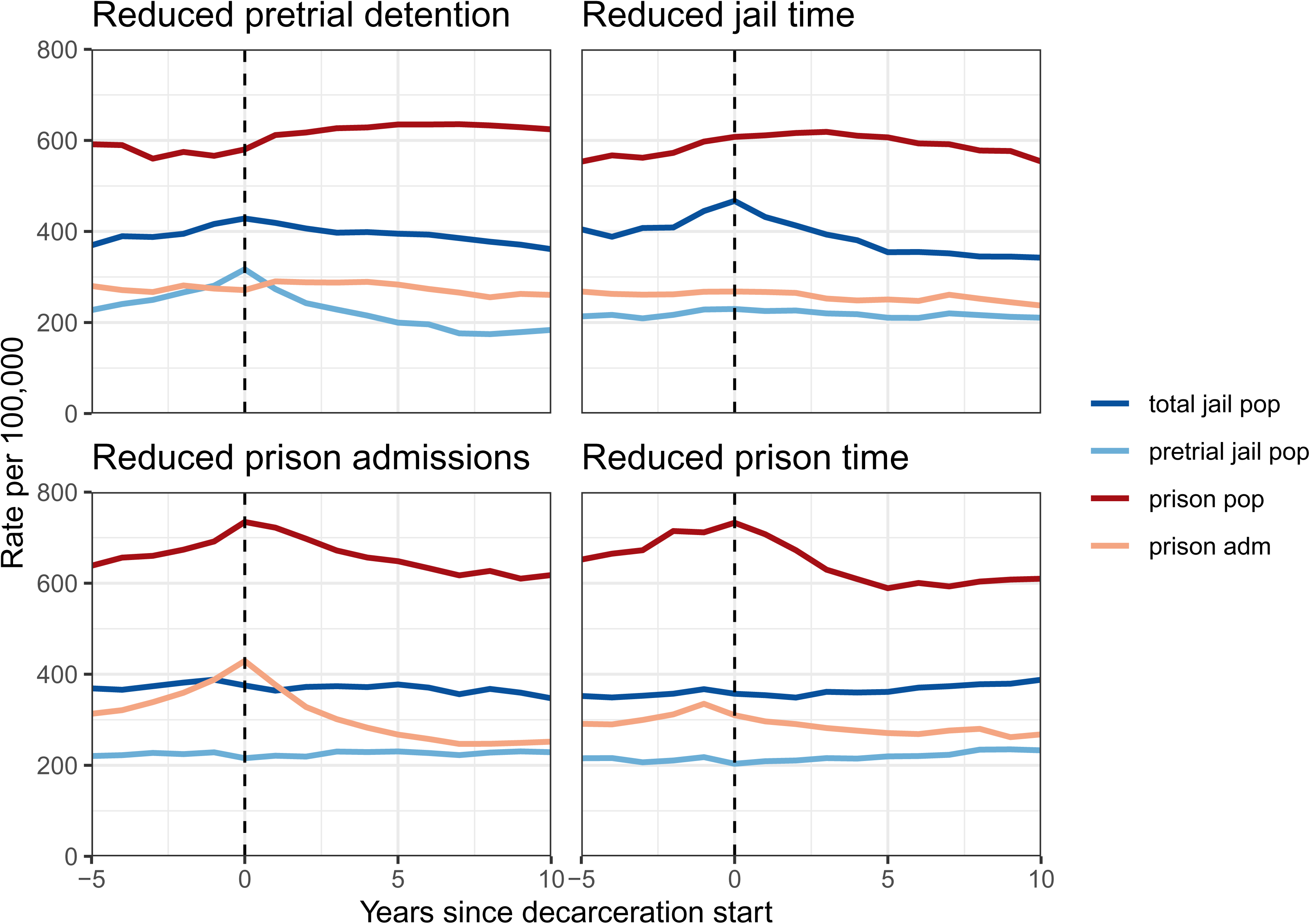
Archetypal trajectories of incarceration measures before and after decarceration onset. Panels show, for each decarceration type, median county-level rates of the four incarceration measures (total jail population, pretrial jail population, prison population, and prison admissions), aligned to the first year a county was classified as experiencing the specified decarceration type (year 0). Each panel includes counties that had at least one episode of that decarceration type during the study period; counties could contribute to multiple panels. Only counties with both jail and prison incarceration data are shown.

Furthermore, counties experiencing each decarceration type tended to have higher levels of the target measure at onset than counties that did not decarcerate or were not yet decarcerating during the study period (**Figure 3, Figure S10**).

The archetypes also reveal potential interactions between jail and prison incarceration. Prison decarceration was not typically accompanied by substantial changes in jail incarceration, but jail decarceration occurred amid slight increases in prison population rates in the years pre- and post-onset.

### Association of decarceration types with mortality

Of the four decarceration types, only reduced pretrial detention was associated with lower mortality overall, with an incidence rate ratio (IRR) of 0.981 (95% CI, 0.969-0.994; 1.9% lower mortality) (**Table 2**). Overall estimates for reduced jail time, reduced prison admissions, and reduced prison time were consistent with the null (IRRs between 0.994-1.007; CIs including 1). Results were similar when all four decarceration types were modeled simultaneously (**Table 2**).

**Table 2.**
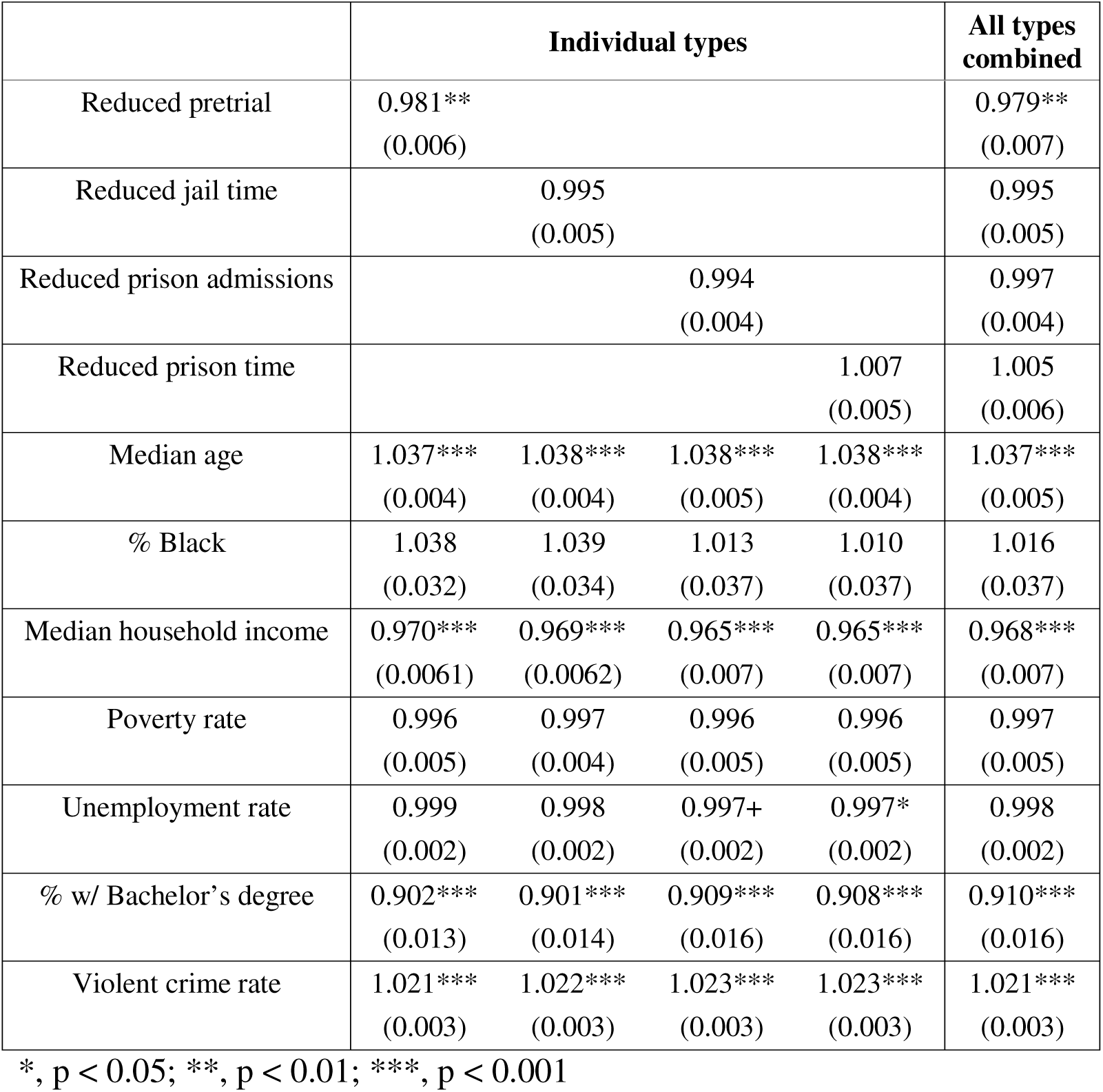
County-level association between decarceration types and all-cause mortality. Results are shown from models including one decarceration type at a time and from a sensitivity analysis including all types in one model. Values are incidence rate ratios (standard errors) for the all-cause mortality rate, adjusted for listed covariates and county and year fixed effects. Median age represents the median across 20 age groups. All other covariates were scaled, so estimates reflect the change associated with a 1 standard deviation shift in the covariates.

These overall estimates, however, obscured significant effect modification by county urbanicity (**Table S3, Figure 4**). Specifically, both reduced pretrial detention and reduced prison admissions exhibited magnified associations with lower mortality in large central metropolitan counties that attenuated to the null across the urban to rural continuum. In large central metropolitan counties, the stratum-specific IRR was 0.966 (95% CI, 0.939-0.993; 3.4% lower mortality) for reduced pretrial detention and 0.981 (95% CI, 0.965-0.998; 1.9% lower mortality) for reduced prison admissions – the latter despite a null overall estimate. In small-midsize metropolitan and rural counties, estimates for both types were near or marginally above 1 (CIs containing 1) (ratios of IRRs with large central metropolitan as reference, 1.024-1.042, p<0.03) (**Table S3, Figure 4**). Associations did not differ significantly by urbanicity for reduced jail time or reduced prison time (**Table S3**).

**Figure 4.**
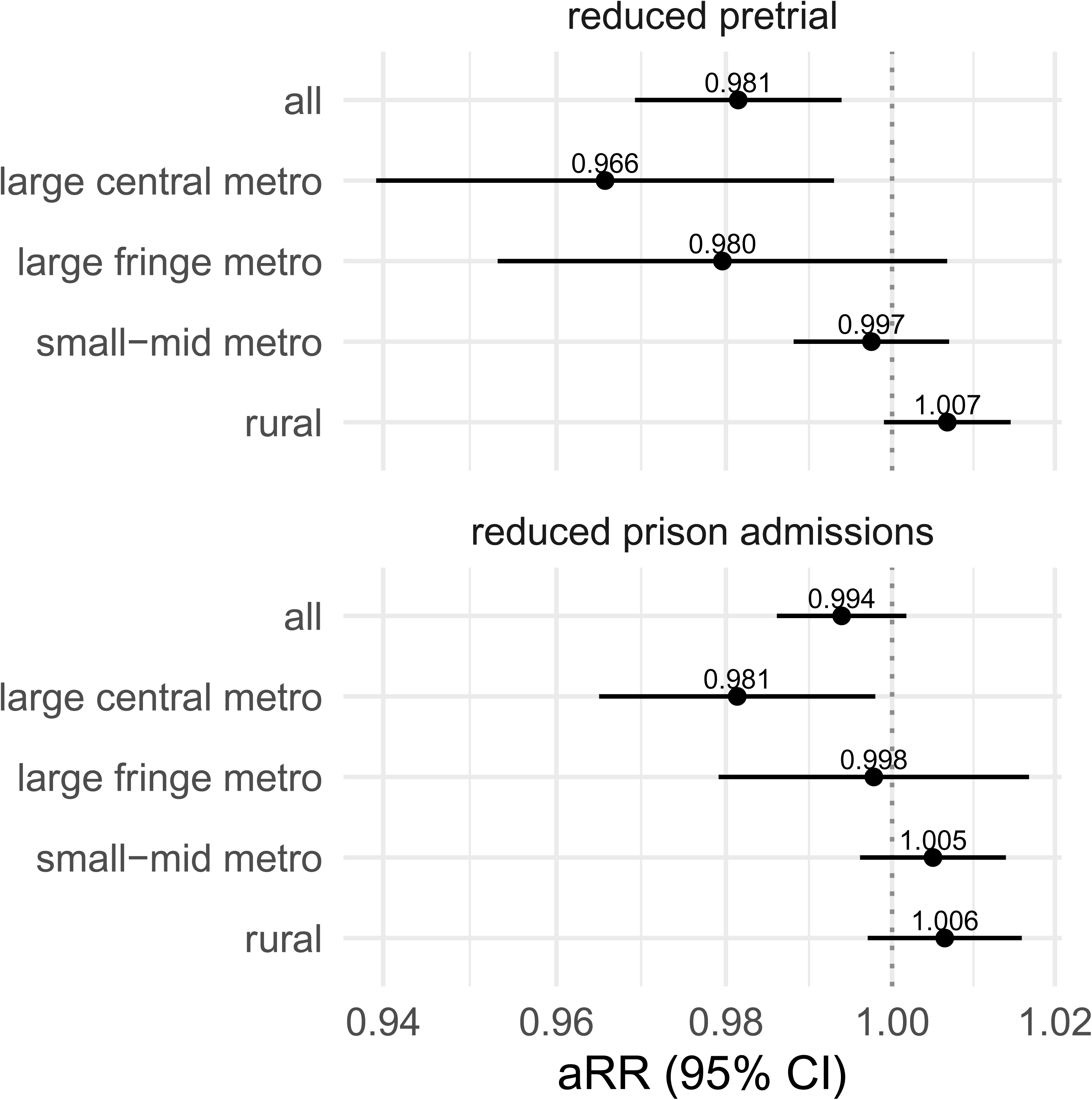
Effect modification by county urbanicity for associations of decarceration types with all-cause mortality. Results are from covariate-adjusted models individually testing each decarceration type, with or without interactions with county urbanicity. Only decarceration types with significant effect modification are shown; all results from this analysis are shown in **Table S3.**

## DISCUSSION

Despite growing interest in decarceration as an opportunity to improve population health, little is known about how decarceration has occurred or whether different approaches to reducing incarceration have different health implications. In this first systematic county-level study of decarceration in the U.S. over two decades, we found that decarceration was common but highly heterogeneous in both its mechanisms and health associations. While most counties experienced at least one form of decarceration, only front-end mechanisms – namely, reduced pretrial detention and reduced prison admissions – were associated with lower mortality, and only in large urban cores. Together, these findings suggest that not all decarceration is equivalent; both the mechanism of decline and the local context in which it occurs appear to shape its population health effects.

A major contribution of this study is demonstrating that the mechanism of decarceration likely matters for health. Prior studies have typically evaluated changes in incarceration rates as though they represent a single phenomenon^10–12,39^. Yet jail populations may decline due to reduced arrests and pretrial detention or shorter jail stays, while prison populations may decline through reduced admissions or shorter sentences. These mechanisms likely act upon different populations and operate through distinct pathways. By developing a reproducible typology using publicly available data, we provide the first systematic characterization of these distinct forms of county-level decarceration, creating a framework for evaluating criminal legal reforms and their health effects according to how incarceration declines, rather than simply whether it declines.

The results suggest that preventing exposure at the initial stages of criminal legal contact could be particularly impactful for community health. Of the four types, only reduced pretrial detention was associated with lower mortality overall. The 1.9% reduction is similar to a prior estimate of a 2.5% change in mortality associated with a quartile change in county jail incarceration^40^, which our findings suggest is driven by pretrial detention. This aligns with literature showing the consequences of even short stints of pretrial detention for employment, housing, family ties, and future justice involvement^41–43^. If short duration of jail exposure can be so harmful, it follows that reducing time in jail may yield marginal gains in comparison, especially if early releases are driven by jail capacity constraints as in several case studies, rather than through purposeful reforms. More research is needed to investigate these varying effects and to examine the specific causes of death that underlie the association between reduced pretrial detention and mortality. Prior studies found no significant effects of cash bail reform in New Jersey on state-level firearm violence or fatal violence against women^13,14^, suggesting other potential pathways through which reduced pretrial detention may reduce mortality.

Our findings also provide new insights into the heterogeneity of prison incarceration and its health implications across local jurisdictions. Although prisons are operated at the state level, the actors that influence who goes to prison and for how long operate at finer geographic levels (e.g. judges, prosecutors)^44^. Yet few studies have broken down prison incarceration across local jurisdictions. In doing so we find considerable local heterogeneity within states following state-level policy reform. Furthermore, we find that reduced prison admissions was associated with lower mortality, at least in large urban cores. Several pathways are plausible: fewer people sent to prison means: 1) fewer people exposed to the health risks of prison, including elevated infectious disease transmission that can spill over to communities^45^; 2) fewer people ultimately released from prison, a transition associated with high mortality risk^46^; and 3) fewer people removed from their families and communities^4,47^. In contrast, reduced prison time showed no significant associations with mortality. As this mechanism acts on the back end to shorten length of stay for people already imprisoned, its effects at the population level may be marginal.

Notably, decarceration’s health effects appear to be context dependent. Beneficial associations were largely concentrated in large urban cores and absent in small-medium metropolitan and rural counties. This is notable in light of smaller cities and rural areas increasingly driving continued carceral expansion in the U.S.^3^. There are a few plausible explanations for this effect modification to test in future research. In places that have historically relied on criminalization and punishment, successful decarceration likely depends on the availability of social services, treatment infrastructure, and other alternatives^48^, which may be more abundant in major urban cores than in smaller cities and rural areas^30,31,49^. Moreover, in small towns, the carceral system may be a source of jobs, commercial activity, and even increased federal and state funds due to prison-based gerrymandering^50^. Smaller economies that have grown dependent on this may be challenged when decarceration is not accompanied by economic transition^51^.

Another possibility is that the drivers of decarceration likely vary across settings. While purposeful reforms may involve simultaneous investments in social and health infrastructure^52^, reactionary decarceration may be more common in resource-constrained areas and less likely to be accompanied by community-based investments. Indeed, most episodes of decarceration involved modest declines confined to a single component of the criminal legal system and frequently followed periods of above-average incarceration, suggesting responses to unsustainable growth rather than deliberate policy transformation. This pattern is consistent with several cases studies and other historical episodes of decarceration that were driven by overcrowding, fiscal constraints, staffing shortages, or litigation^23,53^. In contrast, in large urban cores, decarceration often involved concurrent declines across jail and prison systems, consistent with broader reform strategies such as diversion, decriminalization, or sentencing reform^54^. Future work should examine whether health effects differ according to the policy or practice drivers underlying these distinct mechanisms of decarceration, and to what extent this explains the observed effect heterogeneity across settings.

The atlas enables multiple lines of future research. First, it supports studies examining the association of decarceration types with other health outcomes, particularly those where the mechanism may be particularly consequential. One example is overdose mortality: reduced prison admissions may lower future release volume, while reduced prison time without reduced admissions may increase releases in the short term. Given the established high risk of fatal overdose after prison release^55^, these two mechanisms of prison decarceration could have different effects on community overdose rates. Second, the atlas supports further studies of effect heterogeneity by local contextual factors, such as availability of community-based substance use treatment programs^9,56^. Third, our results highlight substantial within-state heterogeneity in both jail and prison decarceration even following state-level reforms, consistent with the influence of local discretionary actors like judges and prosecutors^57,58^. This creates opportunities to study how local practice mediates the effects of state reforms.

This study has several limitations. Although our analysis included county and year fixed effects and several time-varying covariates, associations with mortality may nonetheless be subject to unmeasured confounding, limiting causal inference. The proposed typology is a first step towards distinguishing mechanisms of decarceration, but categories remain broad. Further distinction was precluded by lack of available data (e.g., direct measures of time served or number of people released to a community). We were unable to identify changes in community supervision due to lack of large-scale county-level data. Given increasing evidence of the harms of community supervision^59,60^, future research should expand to include community-based forms of carceral control. County-level data may contain measurement error, contributing to false positives and negatives. While we attempted to mitigate this, findings should be considered exploratory and hypothesis-generating rather than definitive. Validation was limited to well-documented episodes, typically at the state level or in larger, metropolitan counties. Detection may be lacking in rural counties with smaller populations, where case studies for validation were also limited. Models allowing only up to two breakpoints may oversimplify complex trends, but additional breakpoints risk overfitting given limited observations per county^36^. Trends near the edges of the study period should be interpreted with caution due to systematic exclusion of episodes at the start and end of the period. We did not stratify analyses by gender/sex or race/ethnicity due to substantial missingness in disaggregated data. These are priorities for future research given the disproportionate growth of female incarceration^61^ and prior work suggesting that decarceration may exacerbate racial/ethnic disparities in the criminal legal system^62,63^.

## Conclusion

Between 1999 and 2019, most U.S. counties reduced incarceration through at least one mechanism, although declines were typically modest and confined to one part of the jail or prison system. Front-end mechanisms preventing jail or prison incarceration were associated with lower mortality in large urban counties, but not in smaller metropolitan or rural counties. Mechanisms reducing time served in jail or prison showed no detectable mortality benefits. The findings indicate that the health effects of decarceration depend on both the mechanism of decline and the context in which it occurs. Identifying the conditions for decarceration to improve population health will be essential, particularly in smaller cities and rural areas that increasingly contribute to carceral expansion.

## Data Availability

All data and code referenced or generated in the present work are available at the links below.

https://github.com/yiraneliu/DecarcerationAtlas

https://github.com/vera-institute/incarceration-trends

https://www.cdc.gov/nchs/nvss/nvss-restricted-data.htm

## Acknowledgments

The authors thank Tracey Meares, Caroline Nobo, and Atheendar Venkataramani for helpful discussions.

