## Supplementary Appendix for "A national atlas of county-level decarceration mechanisms and their association with mortality across the United States"

**Supplemental Text**

I. Details on data sources and pre-processing.
II. Diagnosing risk of bias from the two-way fixed effects (TWFE) estimator.
III. Validation details.

**Supplemental Tables**

Table S1. Mapping trends to calendar years and identifying decarceration types in an example county.
Table S2. Sociodemographic characteristics of counties included in jail and/or prison analyses.
Table S3. Associations between decarceration types and mortality by county urbanicity.

**Supplemental Figures**

Figure S1. Piecewise models fit to incarceration measures in an example county.
Figure S2. Map of counties included in analyses of jail and prison incarceration.
Figure S3. Case study validation.
Figure S4. Relative frequency of each decarceration type by state.
Figure S5. Map of jail decarceration types across U.S. counties, 1999-2019.
Figure S6. Map of prison decarceration types across U.S. counties, 1999-2019.
Figure S7. Frequency and timing of combinations of decarceration types, by county urbanicity.
Figure S8. Relative change in target incarceration measures for each decarceration type over time.
Figure S9. Number of counties included in median rates of incarceration measures for archetypal trajectories, by year relative to decarceration onset.
Figure S10. Incarceration rates in decarcerating counties compared to not-yet or never-decarcerating counties.

### I. Details on data sources and pre-processing.

We used the Vera Institute's May 2025 release of the Incarceration Trends Dataset[1]. Data sources used to create this dataset are detailed in Vera's Codebook[2]. Although quarterly jail population estimates were available in recent years for select jurisdictions, we restricted all analyses to annual data to ensure consistent trend ascertainment across counties. When multiple quarterly observations were available within a year, we used quarter two estimates measured at the end of June, consistent with the Bureau of Justice Statistics' standard reporting period. When quarter two data were unavailable, we used the earliest available quarterly observation for that year.

**Table S2** summarizes the characteristics of counties included in the analyses. Counties were included in analyses of jail decarceration if sufficient data were available for both jail measures (pretrial jail population, total jail population). Counties were included in analyses of prison decarceration if sufficient data were available for both prison measures (prison admissions, prison population). Analyses of the frequency and timing of decarceration types (including stratified analyses by urbanicity and state) were restricted to counties with sufficient jail and prison data, to avoid misclassifying missing data as an absence of decarceration.
For analyses involving rates of incarceration measures (e.g. magnitude and archetypal trajectory analyses), we filled in missing data (gaps of no more than three years) using linear interpolation.

Covariate data were sourced and processed as follows. Demographic data on age (median across 20 age groups) and race (percent Black residents) were obtained from the Census via NCI SEER. Poverty rates and median household income were obtained from the Census Small Area Income and Poverty Estimates (SAIPE) program. Educational attainment (percent of the population aged 25 and older with a bachelor's degree) was obtained from the Census via IPUMS NHGIS; values were available for the 1990 and 2000 decennial censuses and for 2008-2018 via American Community Survey (ACS) 5-year estimates, with each 5-year estimate attributed to its middle year (e.g., the 2006-2010 estimate assigned to 2008), and values for 1999 and 2001-2007 obtained via linear interpolation. Unemployment rates were obtained from the Bureau of Labor Statistics' Local Area Unemployment Statistics (LAUS) program via the USDA Economic Research Service and were available annually from 2000-2018, with 2000 values carried backward for 1999. Violent crime rates were obtained from the FBI's Uniform Crime Reporting (UCR) county-level data on reported crimes, downloaded from ICPSR; these were available annually for 1999-2014 and 2016, with 2015 values imputed and 2016 values carried forward for 2017-2018.

### II. Diagnosing risk of bias from the TWFE estimator.

Recent literature has shown that when an exposure of interest occurs at different times across units, its coefficient from TWFE regression represents a weighted sum of estimates from different units and periods, with some receiving negative weights. This can lead to biased estimates when treatment effects are heterogeneous across units or over time[3,4]. Diagnostics for these weights are available for linear but not Poisson models, so we computed a proxy diagnostic using the *twowayfeweights* package in R[5] applied to analogous linear models of the crude mortality rate, adjusted for the same covariates. For each of the four decarceration types, the percent of treated county-years receiving negative weights was low (5-8%). Where all weights sum to 1 by construction, negative weights summed to between -0.002 and -0.012. These results suggest minimal contribution of negative weights to coefficient estimates.

### III. Validation details.

1) **California's Public Safety Realignment**
   a) **Description**: In 2011, California passed two bills collectively known as Public Safety Realignment (AB109 and AB117), in response to a Supreme Court ruling (Brown v. Plata) mandating the state to reduce severe prison overcrowding due to violation of the Eighth Amendment. Realignment shifted responsibility for many individuals convicted of lower-level offenses from the state prison system to county jails. Statewide prison admissions fell by 41% in the first eight months following implementation[6].
   b) **Validation:** Consistent with this documented statewide decline, our approach classified 50 out of 57 California counties as experiencing reduced prison admissions in 2011 (**Figure S3A**). The seven counties not classified as decarcerating during this period had declines that were relatively modes t and/or small in absolute magnitude, and/or noisier data that led to larger standard errors (**Figure S3B**).
2) **Justice Reinvestment in North Carolina**
   a) **Description:** In 2009, North Carolina state leadership sought out support from the Council of State Governments (CSG) Justice Center to identify ways to curb carceral expenditures[7]. In 2011, the recommendations were incorporated into the Justice Reinvestment Act (House Bill 642)**,** a legislative package that reduced the prison population through expansion of drug diversion programs, sentencing reforms, and a shift in housing individuals serving misdemeanor sentences from state prisons to local jails[7]. At the same time, the Act expanded post-release supervision, mandating it for individuals with felony convictions, and introduced new short incarceration sanctions (90-day detention) in response to supervision violations[8]. While these short incarceration sanctions were intended to be a less costly "alternative" to revocation, they resulted in an increased volume of short-term prison admissions for supervision violations. This represents a textbook case of what would be classified as the decarceration type "reduced prison time": where the prison population decreases due to a decrease in the average time served in prison, even while prison admissions remains stable or even increases.
   b) **Validation:** Between 2010-2012, we identified a shift in the predominant type of prison decarceration in North Carolina consistent with the implementation of the Justice Reinvestment Act. Specifically, the number of counties classified as having reduced prison admissions fell sharply, while the number of counties classified as having reduced prison time grew and remained elevated for the rest of the period (**Figure S3A**). Although only a subset of counties was classified as having reduced prison time from 2011 onward, this aligned with underlying empirical trends: counties without reduced prison time experienced rising prison admissions and prison population rates, whereas counties classified as having reduced prison time exhibited declining prison population rates while prison admissions grew (**Figure S3B**).
3) **Voluntary and presumptive sentencing guidelines in Alabama**
   a) **Description**: In response to prison overcrowding, Alabama established the Alabama Sentencing Commission in 2000 and implemented voluntary sentencing guidelines in 2006[9]. The guidelines provided structured recommendations for prison versus non-prison disposition and sentence length, with the goal of reducing prison admissions by diverting individuals with non-violent offenses to non-prison alternatives. This coincided with the expansion of community corrections programs, drug courts, and supervision-based alternatives, making non-prison sentences feasible in select counties[10]. Then, in 2013, Alabama enacted presumptive sentencing guidelines for non-violent offenses, which legally constrained sentencing decisions by establishing default dispositions and sentence ranges[11]. This reform reduced judicial discretion and extended structured

sentencing statewide, including to counties that had not previously adopted the voluntary standards. A prior study found that the presumptive sentencing guidelines reduced average sentence length by almost two years, while reducing race-based and inter-judge sentencing disparities[12].

b) **Validation:** Consistent with this sequence of events, we observed an increase in the number of Alabama counties experiencing reduced prison admissions beginning around 2006, with the number of counties classified as reducing admissions rising from two in 2005 to 20 in 2012 (**Figure S3A**). We observed a subsequent increase in the number of counties classified as experiencing reduced prison time, from 14 in 2012 to 23 in 2017, consistent with the implementation of presumptive sentencing guidelines. Importantly, some counties were classified as not experiencing prison decarceration despite statewide reforms; examination of underlying prison measures in confirms that prison admissions and prison population rates continued to grow in these counties over this period (**Figure S10B**).

**4) Sentencing reforms, expedited releases, and reduced prison admissions in Michigan**

a) **Description:** Starting in the early 2000s, Michigan passed reforms to repeal mandatory minimum sentencing laws and reduce sanctions for property crimes[13]. In 2003, the state also launched the Michigan Prisoner Reentry Initiative (MPRI), focused on expanding and expediting prison releases through expansion of parole board capacity, improved reentry planning, and investments in community-based treatment and reentry infrastructure[13]. State data show a drop in the average minimum length of stay between 2001-2009 and a steady increase in the parole approval rate from 47% in 2000 to 72% in 2017[14], estimated to account for approximately half of the state's prison population decline. Prison admissions also fell during this period due to a combination of factors: declining crime and arrests, expanded diversion programs, and implementation of graduated sanctions and alternatives to revocation for parole and probation violations[13].

b) **Validation:** Our approach identified a surge in the number of Michigan counties experiencing reduced prison time starting in the early 2000s through 2009 (**Figure S3A**). Meanwhile, the number of counties classified as experiencing reduced prison admissions grew slowly but steadily over the study period. Yet, a substantial number of counties were classified as not experiencing either type of prison decarceration during this period. Indeed, prison measures continued to climb until about 2015 in counties labeled as non-decarcerating (**Figure S3B**).

**5) Fiscal pressures, de facto shifts in practice, and pretrial reform in Broward County, Florida**

a) **Description:** After decades of growth, Broward County's jail population began decreasing in 2008, in large part due to financial pressures from the recession and several recent state-level property tax cuts. As tax income fell, county jail budgets were cut, and jail wings closed one after another[15]. As jail capacity decreased, to mitigate overcrowding, police began shifting away from booking individuals for minor offenses, instead giving notices to appear in court[16]. The county also passed an ordinance in 2008 to expand the pretrial release program, reducing the number of people held in pretrial detention for low-level offenses[17]. That same year, the county implemented a pretrial risk assessment tool, COMPAS, which was found to have reduced confinement overall, although it increased racial disparities[18].

b) **Validation:** Our approach identified reduced jail time and reduced pretrial detention in Broward County beginning in 2007 and 2008, respectively, with reduced pretrial detention lasting through the end of the study period (**Figure S3C**).

**6) De facto deprosecution and capacity-based jail releases in Lane County, Oregon**

a) **Description:** Reports from Lane County's Public Safety Coordinating Council document county budget shortages since the early 2000s that resulted in reduced funding of jail

beds and severe understaffing of law enforcement officers, prosecutors, and parole and probation officers[19]. This led to de facto deprosecution of many non-violent misdemeanors and high rates of capacity-based releases (early releases due to overcrowding)[19]. This continued until 2013, when Lane County voters passed a public safety levy to increase the number of funded jail beds, which substantially decreased the number of capacity-based releases, and which has been renewed every five years since[20].
   b) **Validation:** Our approach identified Lane County as experiencing reduced jail time from 2002 until 2013 (**Figure S3C**).

7) **Methamphetamine epidemic, drug court, and precursor laws in Coles County, Illinois**
   a) **Description:** Coles County experienced an acute methamphetamine epidemic in the early 2000s, leading Illinois in meth-related arrests by 2001[21]. In response, local stakeholders formed the Coles County Meth Awareness Coalition in 2003, followed by the establishment of the Coles County Drug Court in 2004, providing treatment-based diversion for individuals charged with drug-related offenses[22]. Additionally, Illinois enacted the Methamphetamine Precursor Control Act in 2005, which sharply restricted access to meth precursors and which were associated with a rapid decline in meth lab seizures across rural Illinois[23].
   b) **Validation:** Our approach identified Coles County as experiencing reduced pretrial detention from 2003 through the end of the study period, aligning with the implementation of the drug court in 2004 and the passage of the meth precursor law in following year (**Figure S3C**). This preceded the peak year of the jail population in 2005 but was consistent with the observed decline in the pretrial jail population through the rest of the period.

8) **Jail decarceration in Orleans Parish and St. Bernard, Louisiana**
   a) **Description:** The jail incarceration rate in Orleans Parish fell by more than 75% during the study period, declining from over 2000 per 100,000 in 1999 to less than 500 per 100,000 in 2019. This decline has been attributed to several local changes: 1) disruptions triggered by Hurricane Katrina and press coverage that drew negative attention to the jail, 2) a series of municipal ordinances in the city of New Orleans encouraging court summons instead of arrests for minor offenses, 3) local reform organizations that blocked construction of a new massive jail complex to replace the flood-damaged facilities, 4) consent decrees between the Department of Justice and both the Orleans Parish Sheriff's Office and the New Orleans Police Department following reports of widespread constitutional violations, 5) introduction of a new pretrial services program to reduce pretrial detention, 6) the ending of a funding system that provided financial incentives for increased incarceration[24].
   b) **Validation:** Our approach identified both types of jail decarceration (reduced jail time and reduced pretrial detention) in Orleans Parish during the study period. Specifically, our approach classified jail decarceration as being driven by reduced jail time between 1999-2008, by reduced pretrial detention between 2008-2011, and by both jail decarceration types from 2012 through the end of the study period (**Figure S3C**). The timing does lack precision compared to the visual trends, but the identification of both jail decarceration types aligns with documented local changes during this period. In the neighboring St. Bernard Parish, which was also severely damaged by Hurricane Katrina, our approach also identified reduced jail time starting in 2006 after the storm (**Figure S3C**).

**Table S1. Mapping trends to calendar years and identifying decarceration types in an example county.** Segment slopes (exponentiated and transformed to represent percentage change) and trend classifications (decreasing, D; non-decreasing, ND) were mapped onto calendar years. Each year, each of the four decarceration types was determined to be present (1) or absent (0) based on criteria in **Exhibit 1**. Corresponds to **Figure S1**. Adm, admissions; red, reduced.

| *year* | *slope jail pop* | *slope jail pretrial* | *slope prison pop* | *slope prison adm* | *trend jail pop* | *trend jail pretrial* | *trend prison pop* | *trend prison adm* | *red. pre-trial* | *red. jail time* | *red. prison adm* | *red. prison time* |
|---|---|---|---|---|---|---|---|---|---|---|---|---|
| *1999* | 5.4 | 15.2 | 4.4 | -9.1 | ND | ND | ND | ND | 0 | 0 | 0 | 0 |
| *2000* | 5.4 | 15.2 | 4.4 | -9.1 | ND | ND | ND | ND | 0 | 0 | 0 | 0 |
| *2001* | 5.4 | 15.2 | 4.4 | -9.1 | ND | ND | ND | ND | 0 | 0 | 0 | 0 |
| *2002* | 5.4 | 15.2 | 4.4 | -9.1 | ND | ND | ND | ND | 0 | 0 | 0 | 0 |
| *2003* | 5.4 | 15.2 | 4.4 | 9.0 | ND | ND | ND | ND | 0 | 0 | 0 | 0 |
| *2004* | 5.4 | 15.2 | 4.4 | 9.0 | ND | ND | ND | ND | 0 | 0 | 0 | 0 |
| *2005* | 5.4 | 15.2 | 4.4 | 9.0 | ND | ND | ND | ND | 0 | 0 | 0 | 0 |
| *2006* | 5.4 | 15.2 | 4.4 | 9.0 | ND | ND | ND | ND | 0 | 0 | 0 | 0 |
| *2007* | 5.4 | -5.1 | 4.4 | 9.0 | ND | D | ND | ND | 1 | 0 | 0 | 0 |
| *2008* | -6.6 | -5.1 | 4.4 | 9.0 | D | D | ND | ND | 1 | 1 | 0 | 0 |
| *2009* | -6.6 | -5.1 | 4.4 | 9.0 | D | D | ND | ND | 1 | 1 | 0 | 0 |
| *2010* | -6.6 | -5.1 | 0.1 | 9.0 | D | D | ND | ND | 1 | 1 | 0 | 0 |
| *2011* | -6.6 | -5.1 | 0.1 | -3.8 | D | D | ND | D | 1 | 1 | 1 | 0 |
| *2012* | -6.6 | -5.1 | 0.1 | -3.8 | D | D | ND | D | 1 | 1 | 1 | 0 |
| *2013* | -6.6 | -5.1 | 0.1 | -3.8 | D | D | ND | D | 1 | 1 | 1 | 0 |
| *2014* | -6.6 | -5.1 | 0.1 | -3.8 | D | D | ND | D | 1 | 1 | 1 | 0 |
| *2015* | -6.6 | -5.1 | 0.1 | -3.8 | D | D | ND | D | 1 | 1 | 1 | 0 |
| *2016* | -6.6 | -5.1 | 0.1 | -3.8 | D | D | ND | D | 1 | 1 | 1 | 0 |
| *2017* | -6.6 | -5.1 | 0.1 | -3.8 | D | D | ND | D | 1 | 1 | 1 | 0 |
| *2018* | -6.6 | -5.1 | 0.1 | -3.8 | D | D | ND | D | 1 | 1 | 1 | 0 |

**Table S2. Sociodemographic characteristics of counties included in jail and/or prison analyses.** Groups (in columns) are based on whether counties have sufficient jail data, prison data, or both; groups are not mutually exclusive. Mean values and interquartile ranges are shown for all variables unless otherwise indicated. Variables are at the county level (i.e., medians are county medians). Characteristics are from the middle of the study period (2009).

| County-level variable | Jail (N=2663) | Prison (N=2184) | Jail and Prison (N=1977) |
|---|---|---|---|
| Median age | 39.2 (36.5-42.1) | 39.2 (36.7-41.9) | 39.1 (36.6-41.8) |
| Median household income ($, thousands) | 43.7 (36.3-48.6) | 43.7 (36.6-48.6) | 44.0 (36.8-49.1) |
| Percent Black | 9.2 (0.5-11) | 8.0 (0.5-9.4) | 8.0 (0.6-9.5) |
| Poverty rate | 9.3 (7.1-11) | 9.5 (7.4-11.3) | 9.6 (7.5-11.3) |
| Unemployment rate | 39.2 (36.5-42.1) | 39.2 (36.7-41.9) | 39.1 (36.6-41.8) |
| Urbanicity/rurality, N (%) | | | |
| Rural | 1591 (60%) | 1290 (59%) | 1146 (58%) |
| Small-mid metro | 655 (25%) | 551 (25%) | 512 (26%) |
| Large fringe metro | 344 (13%) | 278 (13%) | 266 (13%) |
| Large central metro | 63 (2%) | 55 (3%) | 52 (3%) |
| Region, N (%) | | | |
| Midwest | 863 (33%) | 726 (33%) | 643 (33%) |
| Northeast | 180 (7%) | 174 (8%) | 165 (8%) |
| South | 1268 (48%) | 1030 (47%) | 938 (47%) |
| West | 342 (13%) | 244 (11%) | 230 (12%) |

**Table S3. Associations between decarceration types and mortality by county urbanicity.** Incidence rate ratios (IRRs) and 95% confidence intervals (CIs) are shown for each decarceration type and urbanicity stratum, adjusted for covariates, with additional county and year fixed effects. Ratios of IRRs are the estimates from the interaction of decarceration type with urbanicity, with large central metropolitan as the reference stratum.

| Urbanicity | Stratum-specific IRR (95% CI) | Ratio of IRRs (95% CI) |
|---|---|---|
| **Reduced pretrial detention** | | |
| Large central metropolitan | 0.966 (0.939, 0.993) | Reference |
| Large fringe metropolitan | 0.980 (0.953, 1.007) | 1.014 (0.977, 1.053) |
| Small-medium metropolitan | 0.997 (0.988, 1.007) | 1.033 (1.003, 1.063)* |
| Rural | 1.007 (0.999, 1.015) | 1.042 (1.013, 1.073)** |
| **Reduced jail time** | | |
| Large central metropolitan | 0.989 (0.971, 1.007) | Reference |
| Large fringe metropolitan | 1.003 (0.990, 1.015) | 1.014 (0.992, 1.036) |
| Small-medium metropolitan | 0.997 (0.988, 1.006) | 1.008 (0.988, 1.028) |
| Rural | 1.008 (0.997, 1.019) | 1.019 (0.998, 1.041) |
| **Reduced prison admissions** | | |
| Large central metropolitan | 0.981 (0.965, 0.998) | Reference |
| Large fringe metropolitan | 0.998 (0.979, 1.017) | 1.017 (0.991, 1.043) |
| Small-medium metropolitan | 1.005 (0.996, 1.014) | 1.024 (1.005, 1.043)* |
| Rural | 1.006 (0.997, 1.016) | 1.026 (1.006, 1.045)** |
| **Reduced prison time** | | |
| Large central metropolitan | 1.009 (0.989, 1.030) | Reference |
| Large fringe metropolitan | 1.018 (1.002, 1.034) | 1.009 (0.983, 1.035) |
| Small-medium metropolitan | 0.999 (0.990, 1.008) | 0.990 (0.969, 1.012) |
| Rural | 1.001 (0.994, 1.008) | 0.992 (0.971, 1.013) |

*, $p < 0.05$; **, $p < 0.01$

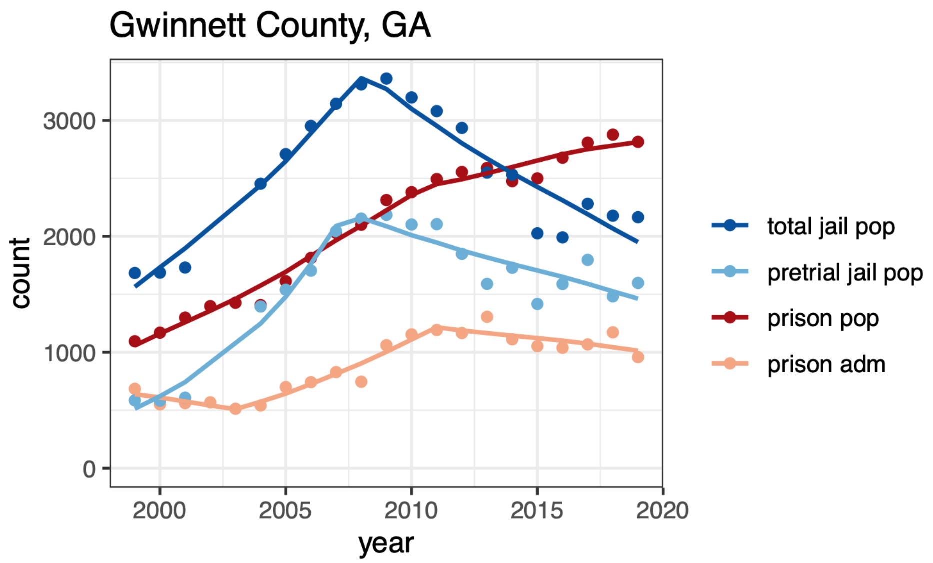


**Figure S1. Piecewise models fit to incarceration measures in an example county.** Points show the underlying data. Lines show the fitted values from piecewise negative binomial regression, conducted separately for each incarceration measure (total jail population, pretrial jail population, prison population, prison admissions). Corresponds to **Table S1**.

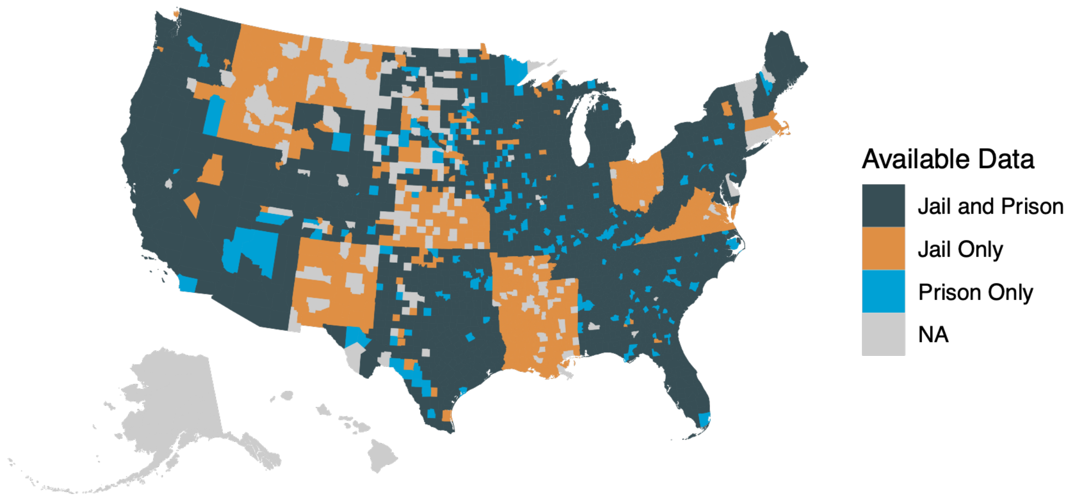


**Figure S2. Map of counties included in analyses of jail and prison incarceration.** NA indicates counties excluded due to insufficient data on jail and prison incarceration.

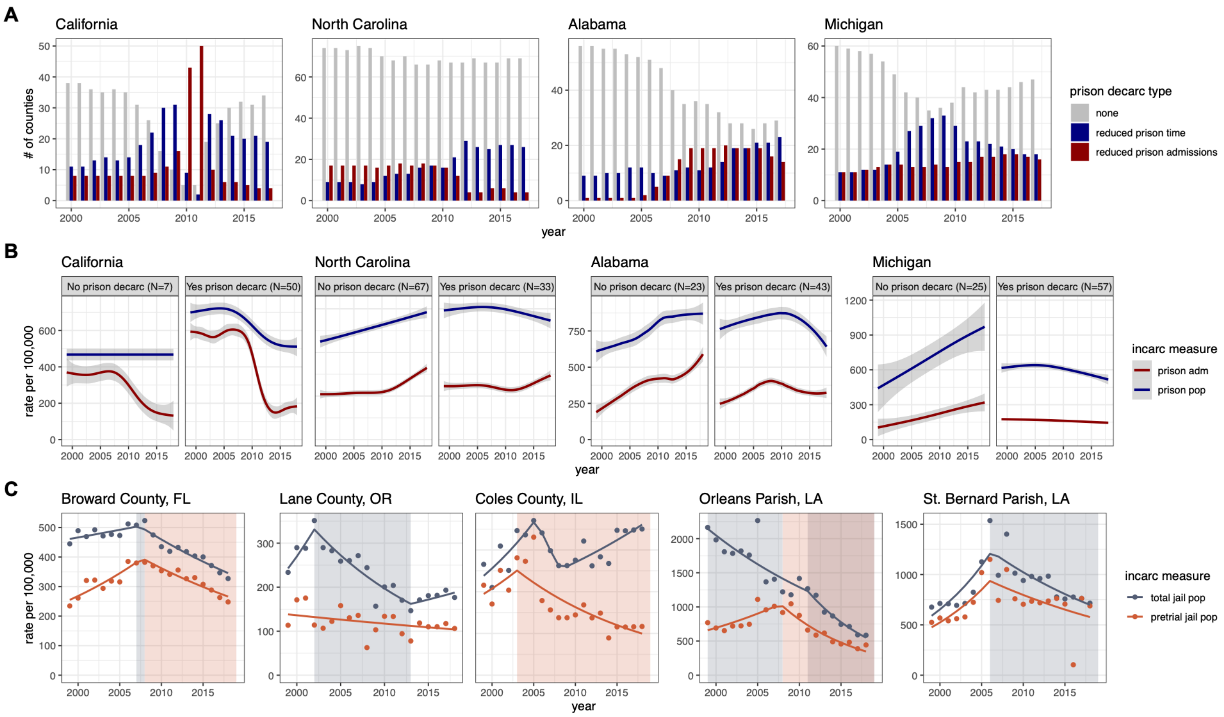


**Figure S3. Case study validation. A)** Bar plots showing the number of counties in each state per year classified as experiencing reduced prison time, reduced prison admissions, or neither. Note: by definition, the two prison decarceration types were mutually exclusive, indicating the primary driver of prison decarceration in a given county in a given year, rather than necessarily representing a complete lack of the other decarceration type. **B)** Locally estimated scatterplot smoothing of trends in the prison admissions rate and prison population rate among counties that were or were not classified as experiencing prison decarceration during the period corresponding to the documented case study. The number of counties in each group is indicated in the facet labels. **C)** Plots of the total jail population rate and pretrial jail population rate in counties that served as case studies for validation of jail decarceration. Points show the underlying data; lines show the piecewise model fit. Periods classified as reduced jail time are shaded in blue-grey, while periods classified as reduced pretrial detention are shaded in orange; brown-grey shading indicates overlapping periods of both types of jail decarceration.

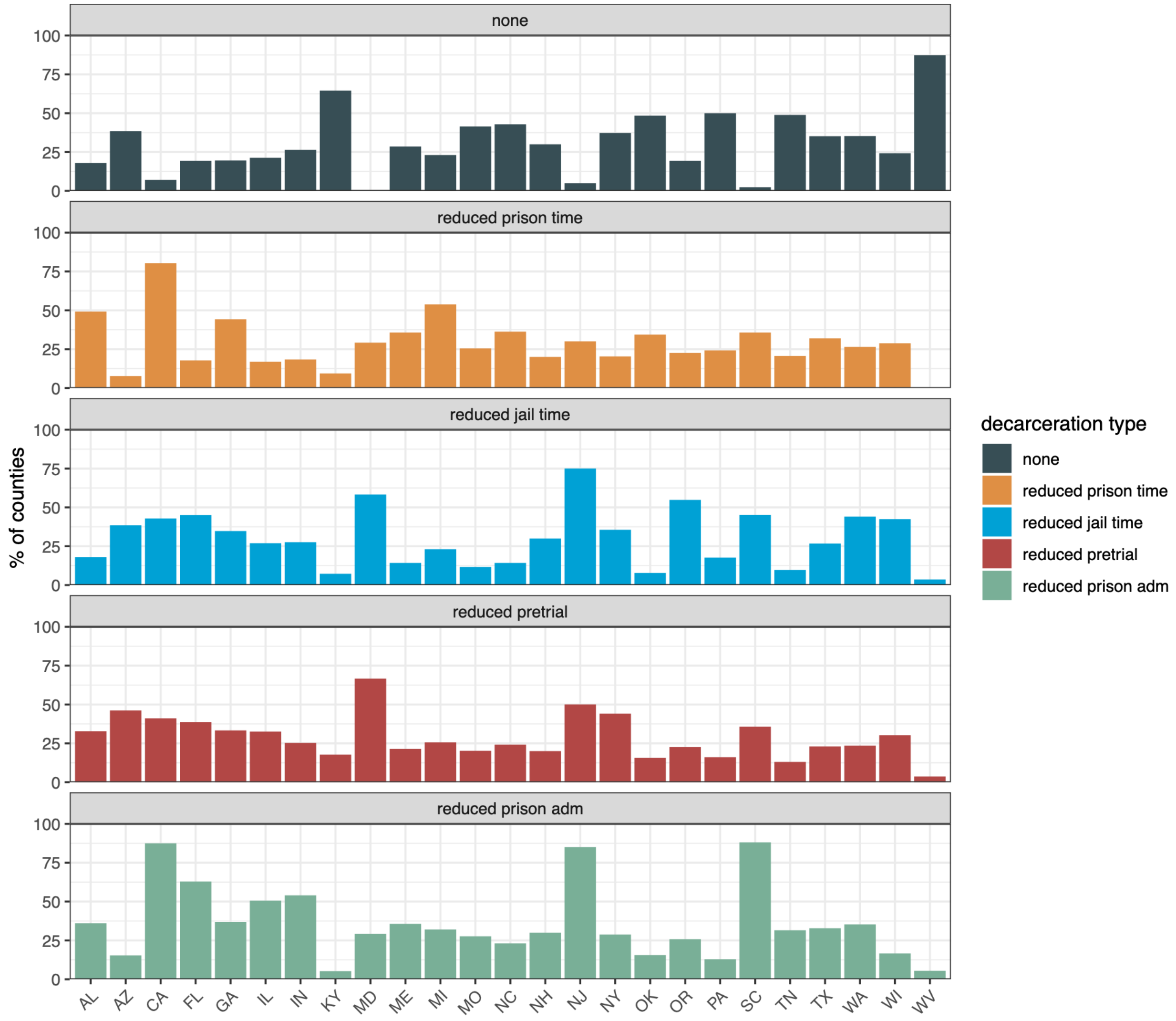


**Figure S4. Relative frequency of each decarceration type by state.** Only states with jail and prison incarceration data for at least 80% of counties are shown.

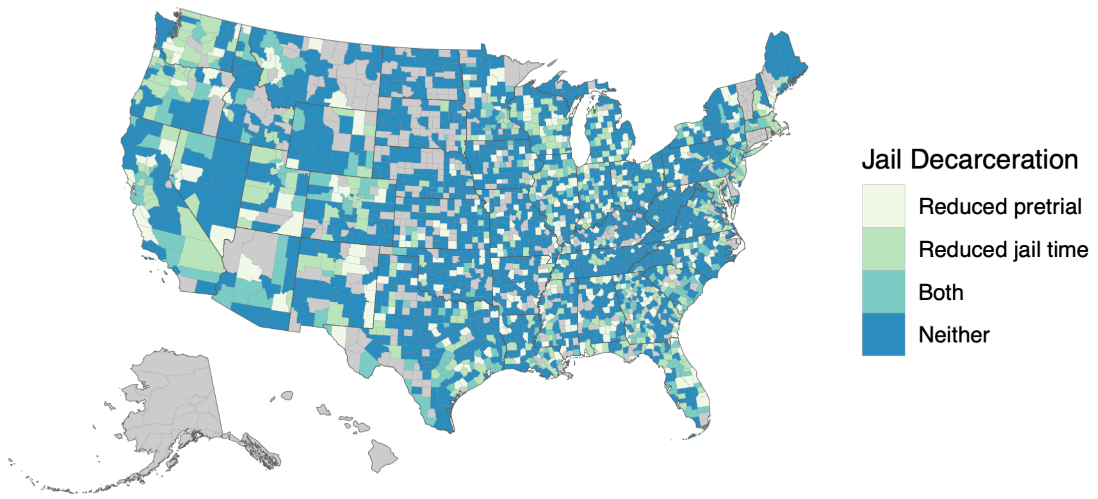


**Figure S5. Map of jail decarceration types across U.S. counties, 1999-2019.** Counties are colored by whether they exhibited reduced pretrial detention, reduced jail time, both, or neither type of jail decarceration during the study period. Counties shaded in grey were excluded from jail analyses due to insufficient data.

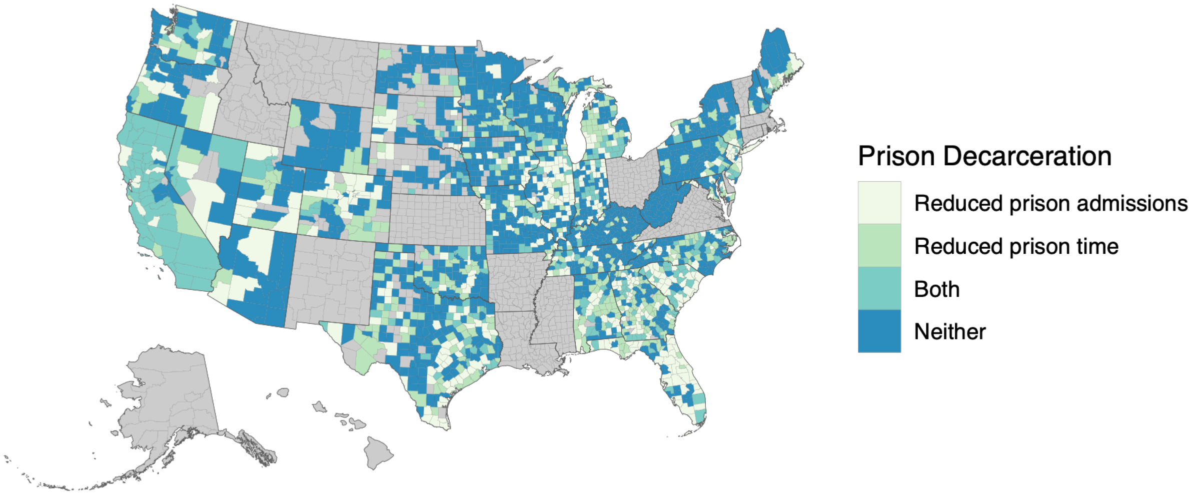


**Figure S6. Map of prison decarceration types across U.S. counties, 1999-2019.** Counties are colored by whether they exhibited reduced prison admissions, reduced prison time, both, or neither type of prison decarceration during the study period. Counties shaded in grey were excluded from prison analyses due to insufficient data.

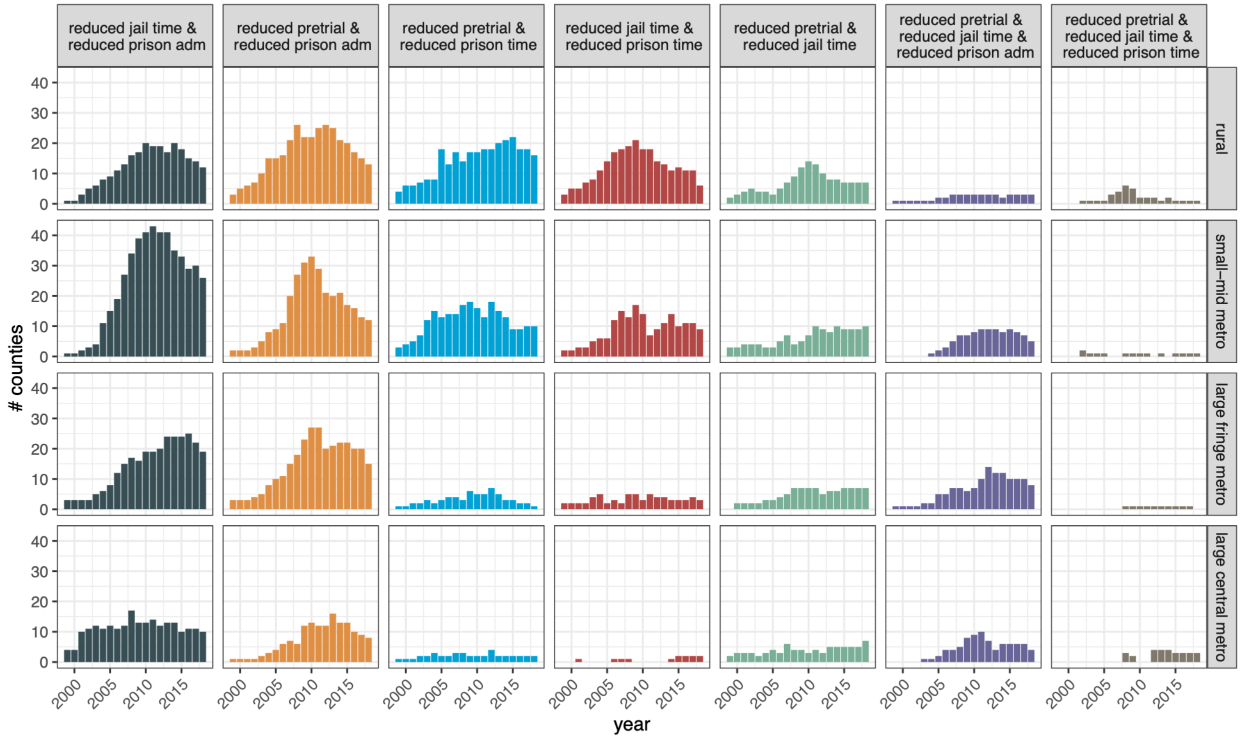


**Figure S7. Frequency and timing of combinations of most common decarceration types, by county urbanicity.** Only counties with both jail and prison data are shown.

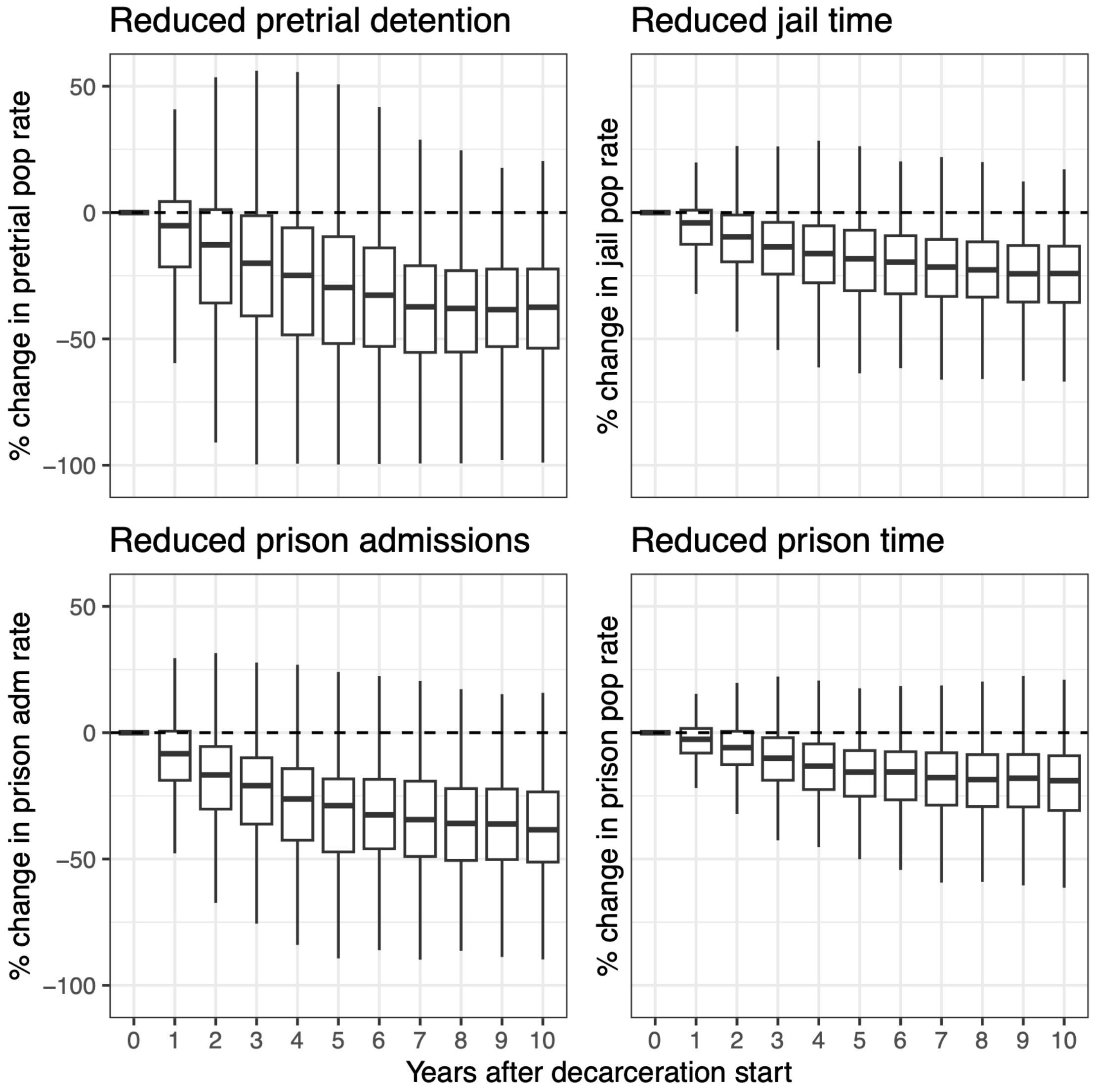


**Figure S8. Relative change in target incarceration measures for each decarceration type over time.** Panels show the percent change in the target incarceration measure corresponding to each decarceration type, relative to the value of that measure at decarceration onset (year 0). Each panel includes counties that had at least one episode of that decarceration type during the study period; counties could contribute to multiple panels.

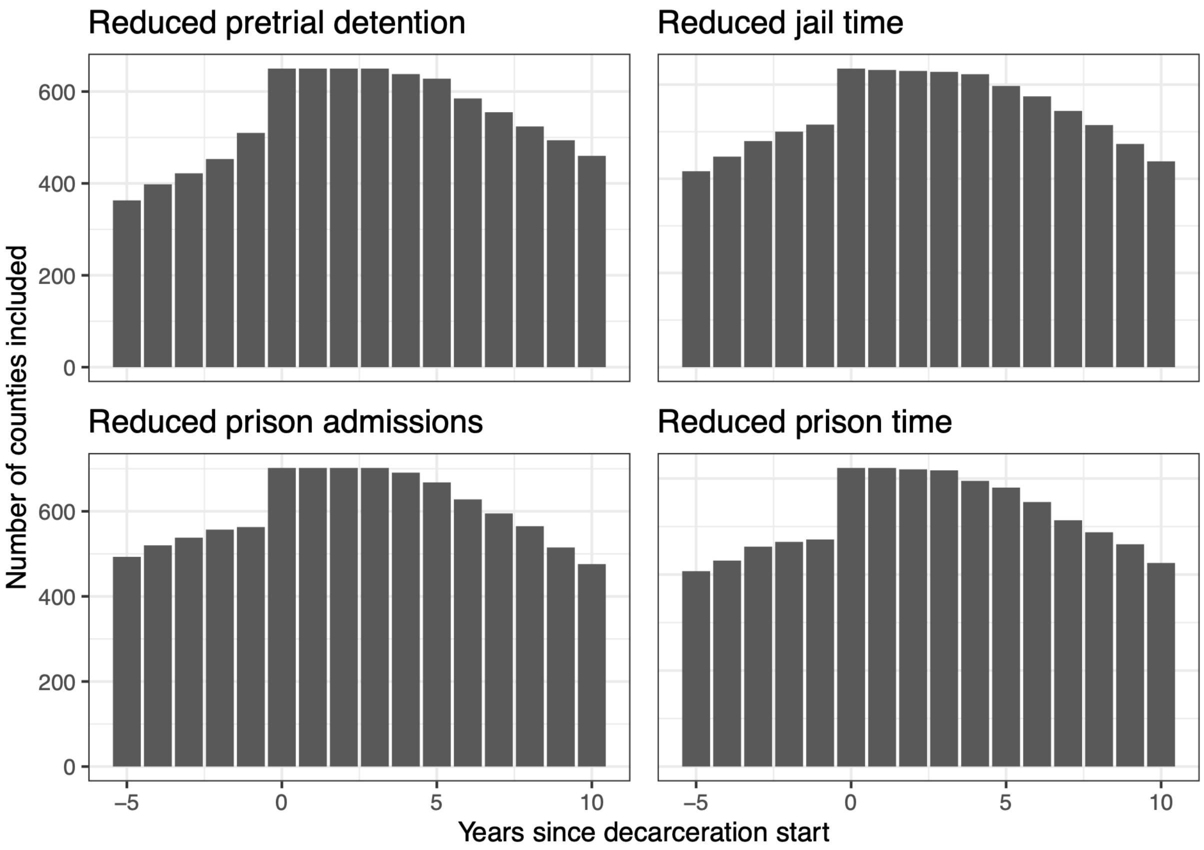


**Figure S9. Number of counties included in median rates of incarceration measures for archetypal trajectories, by year relative to decarceration onset.** Corresponds to Exhibit 4 in the main text, showing the number of counties with each decarceration type that contributed data on the four incarceration measures in each year preceding and following decarceration onset.

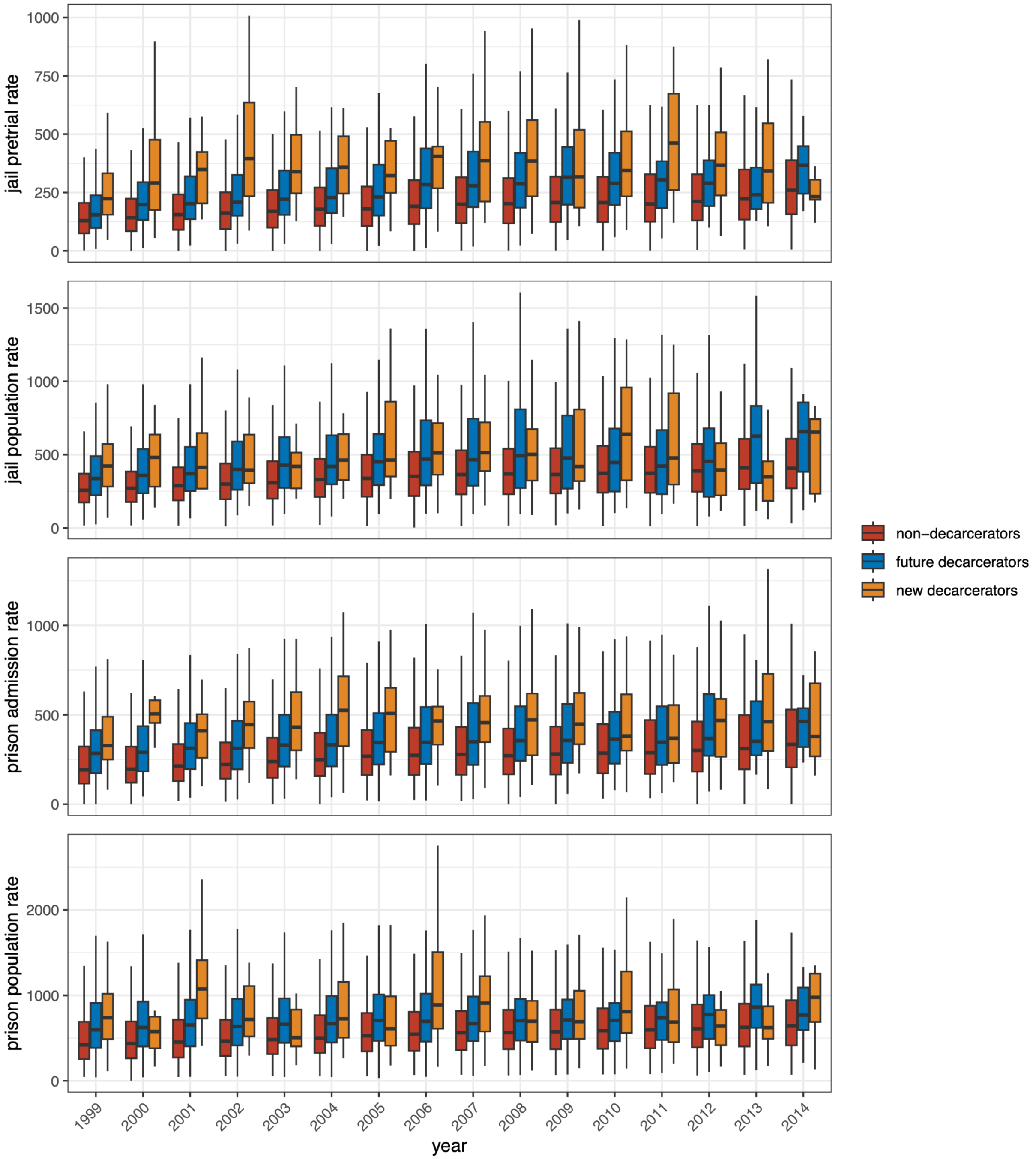


**Figure S10. Incarceration rates in decarcerating counties compared to not-yet or never-decarcerating counties.** Each panel shows a target incarceration measure corresponding to each of four decarceration types: reduced pretrial detention, reduced jail time, reduced prison admissions, and reduced prison time. Boxplots show counties that, in a given year, are starting that type of decarceration ("new decarcerators"), have not yet started but will in a subsequent year ("future decarcerators"), or that do not exhibit that decarceration type throughout the study period. The figure is truncated for 2015 and later due to an imposed condition requiring decarceration to start at least five years prior to the end of the period to avoid spurious edge effects.
